# Type, location and zygosity of *KCNJ16* mutations may determine the clinical severity of Hypokalemic Tubulopathy and Deafness (HkTD)

**DOI:** 10.64898/2026.01.07.25343066

**Authors:** Leire Gondra, Shivani Mora, Imran G Shaikh, Alejandro Garcia-Castaño, Franziska Schilling, Gema Ariceta, Leire García-Suarez, Patricia Tejera-Carreño, Gema Fernández-Juarez, Alfredo Santana Rodríguez, Jonai Pujol-Giménez, Marta García-Alonso, Sara Gómez-Conde, Ainhoa Camille Aranaga-Decori, Martin Koemhoff, Vijay Renigunta, Stefanie Weber, Leire Madariaga, Aparna Renigunta

**Author notes:** **Correspondence:** Dr. Aparna Renigunta, Department of Pediatric Nephrology, University Children’s Hospital, Philipps University Marburg, 35043, Marburg, Hessen, Germany, Dr. Leire Madariaga, Department of Pediatric Nephrology. Cruces University Hospital, University of the Basque Country, Biobizkaia Health Research Institute Plaza de Cruces s/n, 48903, Barakaldo, Bizkaia, Spain. Shared Senior and Correspondence (Leire Madariaga, Aparna Renigunta).

## Abstract

The pivotal role of K_ir_5.1 (*KCNJ16*) in maintaining electrolyte and acid-base homeostasis was demonstrated by animal studies and highlighted by the identification of disease-causing mutations in *KCNJ16* resulting in a complex tubulopathy with variable severity. Although the underlying molecular mechanisms remain elusive, the modus operandi of K_ir_5.1 is rooted in its heteromeric association with K_ir_4.1 (*KCNJ10*) and K_ir_4.2 (*KCNJ15*). The ubiquitous expression of *KCNJ16* and the heterogenous clinical picture point towards the importance of protein-protein interactions and membrane trafficking of the heteromeric channels involving K_ir_5.1. Alongside a retrospective report on the R137C variant, we describe a divergent clinical phenotype in a patient compound heterozygous for I132R and R176*. Moreover, we characterize a novel variant (homozygous for K48* and Y57*) whose clinical presentation coincides with the phenotype of the *Kcnj16^-/-^* mouse model (lack of deafness). Functional studies, buttressed by biomolecular studies (imaging and Co-IPs) in mammalian cells emphasize that the type and location of the *KCNJ16* mutations may determine their interaction, membrane localization, and consequently, channel function and the resulting clinical phenotype. Our study highlights the need for molecular understanding of the *KCNJ16* variants to promote correct diagnosis and personalized therapies, especially those involving channel modulators.

## Introduction

Inward rectifying potassium (K_ir_) channels encoded by *KCNJ* genes, are broadly expressed in human cells and are fundamental to the regulation of resting membrane potential, electrolyte equilibrium and cellular excitability [1]. As the name suggests K_ir_ channels uniquely enhance conductance upon hyperpolarization and reduce it upon depolarization stabilizing the resting membrane voltage of non-excitable cells near the K^+^ equilibrium potential [2]. In the kidney, K_ir_ channels are an integral part of the polarized epithelia in renal tubules and play an essential role in enabling transport processes across the apical and basolateral membrane.

Pathogenic mutations in *KCNJ1* encoding homomeric potassium channels K_ir_1.1 (ROMK), cause Bartter’s syndrome type II (BS-II), marked by polyhydramnios and profound salt and fluid loss both pre- and postnatally. The etiology of BS-II lies in reduced potassium recycling, resulting in defective reabsorption of sodium chloride in the thick ascending limb of Henlès loop (TAL) [3, 4]. Unlike the apical ROMK channels, basolateral K_ir_ channels of the distal convoluted tubules (DCT) (K_ir_4.1 encoded by *KCNJ10*) and proximal tubules (PT) (K_ir_4.2 encoded by *KCNJ15*) heteromerize with their close homologue K_ir_5.1 (encoded by *KCNJ16*) at the basolateral membranes and partake in the potassium recycling in these tubular segments. This heteromerization modulates the properties (rectification, sensitivity to pH_i_) of the resulting channels [5–7]. Although the vital role of the inherently functional basolateral potassium channels K_ir_4.1 and K_ir_4.2 has been underlined by studies involving knockout animals, disease-causing variants in *KCNJ15* affecting renal function have not been so far reported [8]. Juxtaposing this, the essential role of basolateral potassium recycling for transepithelial salt reabsorption was particularly highlighted by the discovery of recessive loss-of-function variants in *KCNJ10* causing EAST/SeSAME syndrome [9, 10]. These patients have a Gitelman-like phenotype due to reduced activity of the sodium-chloride co-transporter (NCC) in the DCT, resulting from a decrease of potassium recycling at the basolateral membrane through defective homo and heteromeric channels comprising K_ir_4.1 and K_ir_4.1/K_ir_5.1 respectively. The clinical picture of EAST/SeSAME patients features salt wasting, RAAS activation, hypokalemic metabolic alkalosis, hypomagnesemia, and hypocalciuria. In addition, they exhibit sensorineural deafness, ataxia, and intellectual disability due to K_ir_4.1 dysfunction in the inner ear and the central nervous system.

Recently, disease-causing variants in *KCNJ16* have been added to the continuously evolving spectrum of renal tubular salt wasting disorders [1, 11]. *KCNJ16* is expressed in various organs and tissues, such as the heart, brain, inner ear, kidney, pancreas, thyroid, and testis, where its expression coincides with that of other *KCNJ*s [5]. Interestingly, K_ir_5.1 does not form functional channels alone. However, in brain neurons, the C-terminus of K_ir_5.1 interacts with PSD-95 (Post Synaptic Density protein-95) to form functional channels responsible for synaptic transmission [12]. In the kidney, K_ir_5.1 heteromerize with K_ir_4.1 and K_ir_4.2 to form functional channels. *KCNJ16* variants result in a complex, pleiotropic, monogenic tubulopathy sometimes referred to as HkTD (Hypokalemic Tubulopathy and Deafness) characterized by hypokalemia, salt wasting, varying severity of acidosis and sensorineural deafness [1, 11, 13]. Among the patients so far reported, one patient (T64I) stands out with a renal clinical picture mirroring EAST/SeSAME showing alkalosis [1]. In addition to HkTD, recent studies in humans and animals have demonstrated the multifaceted involvement of these silent channels in various diseases like salt-sensitive hypertension, subfertility, variety of cancers, and impaired respiratory chemoreception causing SIDS (Sudden Infantile Death Syndrome)[14–17].

Given the variegated clinical presentation, the broad distribution of K_ir_5.1 channels and their ability for heteromerization, we hypothesize that molecular characteristics of protein interactions and the membrane localization of the resulting heteromeric channels (involving mutant K_ir_5.1) should be closely examined. Supporting evidence of K_ir_ channel mutations associated with altered channel expression at the plasma membrane resulting from defective protein-protein interaction (with subunits, accessory proteins etc.,) and/or defective channel trafficking is reviewed elsewhere [18]. Furthermore, such molecular understanding is particularly valuable for exploring the scope of K_ir_ channels as pharmacological tools and probing the therapeutic potential of targeted modulators [19]. In this study, we report 4 patients from 3 families with salt-losing tubulopathy secondary to pathogenic biallelic variants in *KCNJ16*. Here, we report a novel nonsense variant Y57* in family C. The index patient is double homozygous for two nonsense variants (Y57* and a previously reported K48* [11]). The two patients in family A are affected by the pore mutant R137C, reiterating the clinical findings reported previously [1]. However, the clinical picture of the patient from family B, who is compound heterozygous for I132R and R176*, differs substantially from that previously reported, further contributing to the growing complexity of the clinical spectrum associated with HkTD. In addition, we have also included the *KCNJ16(T64I)* variant presenting alkalosis instead of acidosis in our molecular analysis. We used a battery of experiments involving electrophysiology, protein and cell biochemistry to demonstrate that the nuanced clinical presentation of HkTD arises from, either the inability to form functional tetramers or varying degree of dominant negative effects of the mutant K_ir_5.1 on both K_ir_4.1 and K_ir_4.2. Thus, the type, location, zygosity and allelic status of the *KCNJ16* variants may determine the clinical picture of the affected individuals.

## Methods

### Patients and Genetic Screening

Ethical approval for this study was obtained from the Ethics Committee for Clinical Research of Euskadi (CEIC-E). Informed consent was obtained from each proband or their legal tutors and their families before inclusion in the study. The research was carried out in accordance with the Declaration of Helsinki on human experimentation of the World Medical Association. Genomic DNA of affected individuals and available family members was extracted from peripheral venous blood by standard methods. The whole exome sequencing was performed on all subjects using the Ion AmpliSeq™ Exome RDY Library Preparation kit followed by sequencing by the Ion GeneStudio S5 Plus (Thermo Fisher Scientific). We used the Ion Reported Software for variant filtering. Variants were filtered to include only the coding and flanking intronic variants (+/-10bp) with a Minor Allele Frequency (MAF) ≤ 0.05 in 1000 Genomes Browser, Exome Aggregation Consortium (ExAC) and ESP Exome Variant Server (ESP). We filtered genes related to HPO (Human Phenotype Ontology) terms used to direct the search for genetic variants relevant to the patient: Hypokalemia-HP_0002900 and Renal salt wasting_HP_0000127. We filtered variants previously described as benign or likely benign in ClinVar database and UCSC common SNPs. Sanger sequencing was performed for variant confirmation. Variants found were validated by polymerase chain reaction (PCR) followed by direct sequencing of both strands (conventional Sanger sequencing) and loaded onto an ABI3500 Genetic Analyzer (Thermo Fisher Scientific, California, USA).

### Molecular Biology

The cDNAs for *KCNJ16* (NM_001270422), *KCNJ10* (NM_002241), and *KCNJ15* (NM_001276435) were cloned in the mammalian expression vectors: pmRFP-C1 (Acc.Nr: AAM54544), pEGFP-C1(Cat#6084-1, AccNr: U55763) (Clontech, Mountain View, USA); in the MCS1 of the modified pIRES (T) plasmid with 3x Flag introduced after Xho1 site of MCS1 (MCS2 without and with EGFP for transfection control) (Acc.Nr: JX088702, Texas, USA), for studies involving mammalian expression; and between the UTR of the Xenopus beta-globin gene in pSGEM or pTLN vectors [1] for studies involving expression in Xenopus *laevis* oocytes. Site-directed mutagenesis was performed using PfuUltra II Hotstart PCR Mastermix (Agilent) to introduce the desired mutations in the *KCNJ16* clones. Complementary RNA (cRNA) was transcribed *in vitro* from linearized plasmids containing the cDNA of interest using T7 or SP6 kit (New England Bio Labs Inc., Frankfurt, Germany & Ambion, Huntingdon, United Kingdom). cRNA was purified by LiCl/ethanol precipitation. Yield and concentration were quantified spectrophotometrically, and the quality of RNA was confirmed by agarose gel electrophoresis.

### Cell culture

HeLa cells (Biozol; Cat#ADD-C0008001) and HEK-293 cells (Cytion; Cat#CLS-300192) were cultured in high-glucose DMEM supplemented with 10% fetal calf serum (FCS; Life Technologies, Paisley, United Kingdom) and 1% penicillin/streptomycin (PAA Laboratories, Pasching, Austria), and were transiently transfected using jet PRIME reagent (Polyplus), according to the manufacturer’s instructions. For live-cell imaging using confocal microscopy, HeLa cells were seeded into 35 mm dishes (Greiner Bio-one GmbH, Frickenhausen, Germany) containing coverslips. Cell lines were obtained from either commercial companies or from the repositories as mentioned and used as received, consistent with their established authentication. All cell lines used were confirmed to be mycoplasma-free.

### Two Electrode Voltage Clamp Measurements

TEVC measurements were performed as described previously [1]. Defolliculated X. *laevis* oocytes (Ecocyte Bioscience, Dortmund, Germany) were injected with 50 nl nuclease-free water containing cRNA (1–2.5 ng/oocyte of *KCNJ10* or *KCNJ15* for single injections; for co-injections *KCNJ16* (wt, variant or wt+variant) and the heteromeric partner were injected at a 1:1 ratio). Oocytes were incubated at 16°C in ND96 solution containing 96mMNaCl, 2 mM KCl, 1 mM MgCl2, 1.8 mM CaCl2, and 5 mM HEPES (pH 7.4) supplemented with 100 mg/ml gentamycin and 2.5 mM sodium pyruvate. Two to three days post-injection, two-electrode voltage-clamp measurements were performed at room temperature using a Gene Clamp 500 amplifier (Axon Instruments) in bath solution containing ND96. Currents were evoked by 200-ms voltage steps from −120 to +40 mV in 20mV increments from a holding potential of +80 mV. Data are presented as mean ±SEM; Statistical analyses were performed using ANOVA. Experiments were repeated in at least three independent oocyte batches.

### Live Cell Imaging

Live cell imaging was performed in HeLa cells as described previously [20]. In brief, HeLa cells were co-transfected with GFP or RFP-tagged *KCNJ* constructs (250 ng) and organelle markers (250 ng). Used are plasmids PLCδPH-pRFPC3 for plasma membrane and pDsRed2-ER for ER staining [21]. Lysoracker Red DND-99 (2µM, Cat#L7528, Thermo Fischer) was used to detect lysosomes. Cells were analyzed 48 hours after transfection. Live cell images were captured using ZEISS 710 LSM upright confocal microscope. EGFP and RFP images were acquired by excitation of two lasers at 488 and 561 wavelengths and emission was measured using 493-556 and 578-696 nm filters, respectively. Live images were acquired and later analyzed using ZEN Black SR 3.0 software.

### Co-immunoprecipitation

Co-immunoprecipitations were performed in HEK293 cells as described previously [22, 23]. HEK293 cells were transfected singly or co-transfected with C-terminus flag-tagged *KCNJ16* (WT or variants T64I, R137C, R176*) and GFP-tagged *KCNJ10* or *KCNJ15*. After 48 h, cells were lysed in Triton X-100 buffer (50 mM Tris-HCl, pH 7.5, 150 mM NaCl, 1% Triton X-100) containing 10µl/mL of protease-Inhibitors. Immunoprecipitation was carried out using anti-Flag M2 magnetic beads (Sigma Aldrich) and GFP-Trap Magnetic Agarose (ChromoTek) for 12 h at 4° C. Immunoprecipitates were washed (3x) with PBS and eluted in 2x SDS loading buffer and boiled at 72° C for 10 min. Eluted proteins were resolved by SDS-PAGE, transferred and probed with rabbit anti-flag antibody (1:1000, Cat# F7425, Merck) and mouse anti-GFP antibody (1:1000, Cat#sc-9996; RRID: AB_627695, Santacruz), followed by fluorescence-conjugated secondary antibody (1:10000, Goat anti-mouse star bright 700_Cat# 12004158 and goat anti-rabbit star bright 520_Cat#12005869, Biorad). Bands were visualized using the Bio-Rad Chemidoc MP imaging system.

### Statistical Analysis

Data was analyzed in graphpad prism 6. Reported are means ± SEM (Standard error of mean). Statistical significance was determined using ANOVA. In the figures, statistically significant differences to control values are marked by *asterisks* (****, *p* < 0.0001); *n.s* indicates non-significant differences (*p* > 0.05).

## Results

### Clinical case description

In this study, we present four patients from three families (family A, B and C in Figure 1) with pathogenic variants in *KCNJ16*, their clinical presentation and their clinical course. Clinical details and genetic findings are shown in Table. 1.

**Fig. 1.**
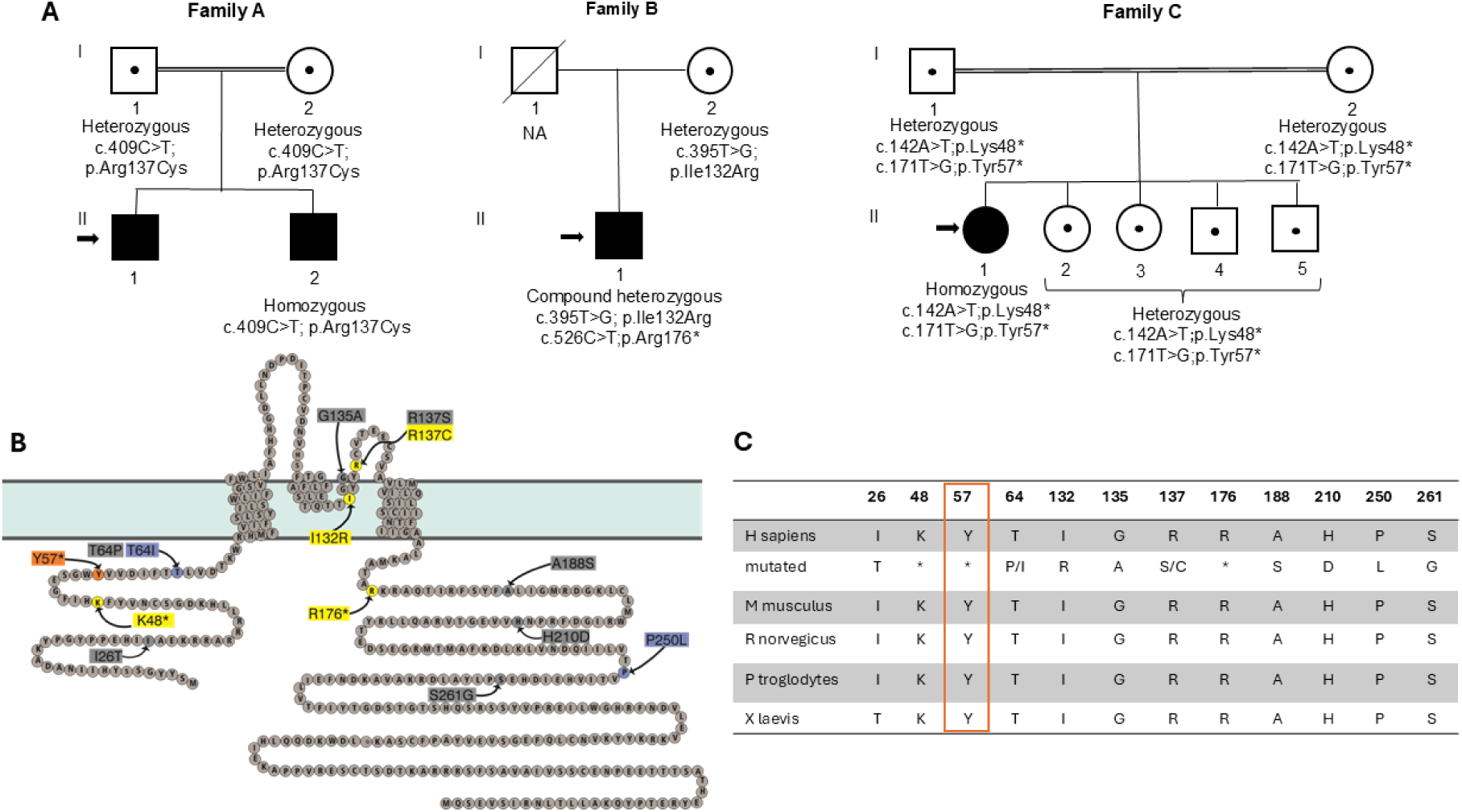
Family pedigrees and localization of identified variants in *KCNJ16* (K_ir_5.1). **(A)** Pedigrees of three families with four affected individuals carrying homozygous or compound heterozygous *KCNJ16* variants. Index cases are indicated by arrows and parental consanguinity by double bars. Zygosity was confirmed by segregation analysis. Zygosity for *KCNJ16* variants was analyzed in the parents as indicated. In Family C, both variants map to the same allele, based on shared NGS reads. NA. not available. **(B)** Variant map of K_ir_5.1 showing all 13 previously reported variants and the novel variant (orange). Other variants from this cohort (yellow), additional study-related variants (blue), and variants not included here (gray). **(C)** Conservation of all reported *KCNJ16* variants across the species; the novel Y57* variant is highlighted in orange.

**Table. 1.** Clinical characteristics and genetic findings of patients in our cohort.

| Variable | A-II-1 | A-II-2 | B-II-1 | C-II-1 |
| --- | --- | --- | --- | --- |
| Sex | Male | Male | Male | Female |
| Consanguinity | Yes | Yes | No | Yes |
| Age at diagnosis | 15 – 19 months | 5 – 9 months | 22 – 26 years | 10 – 14 months |
| Weight/length, SD | -3/0.38 | -1.7/-0.8 | ND/+0.8 | -3/-2.1 |
| Clinical findings | Vomiting, fever, polyuria, polydipsia, salt craving | Vomiting, growth failure, mild polyuria | Asthenia, weakness, cramps, mild polyuria | Growth failure, weakness, Asthenia |
| **Blood at presentation** |  |  |  |  |
| Na<sup>+</sup>, NV 135-145 meq/l | **126** | 133 | 140 | 136 |
| K<sup>+</sup>, NV 3.5-5.5 meq/l | **1.5** | **2.7** | **1.5** | **1.3** |
| Cl<sup>-</sup>, NV 100-109 meq/l | **87** | **95** | 103 | **99** |
| Ca, NV 8.5-10 mg/dl | ND | ND | ND | 9.7 |
| Mg, NV 1.6-2.5 mg/dl | 2.2 | 1.9 | 1.9 | 2.4 |
| Cr mg/dl (eGFR) | 0.28 (122) | 0.35 | **0.99 (63)** | **0.5 (65)** |
| HCO<sub>3</sub><sup>-</sup>, NV 22-26mmol/l | **21** | **17** | **16** | **21** |
| Renin, NV 0.3-1.8ng/ml/h | **>30** | **>30** | ND | **> 45** |
| Aldosterone, NV 2.5-30ng/dl | **>100** | **>100** | ND | **850** (NV 35-300pg/ml) |
| **Urine at presentation** |  |  |  |  |
| pH, NV 5-8 | 6 | 6 | 5.5 | 6 |
| FE-Na, NV <1 % | 0.15 | 0.8 | ND | 0.83 |
| FE-Cl %, NV <1 % | 1.9 | 1.4 | ND | 0.9 |
| FE-K, NV <15 % | **106.5** | **29.7** | **18.3** | **57** |
| TTKG, NV 2-10 | ND | ND | ND | **27.8** |
| Ca/Cr, NV < 0.5 (0-2 y), <0.2 mg/mg (>2 y) | 0.1 | 0.3 | ND | 0.02 |
| **Clinical data at last follow-up** |  |  |  |  |
| Age | <15 years | <10 years | 37 - 41 years | <5 years |
| Weight (SD) | 0.1 | -0.8 | ND | -2 |
| Length (SD) | -0.87 | -1.4 | + 0.8 | -1.7 |
| Acidosis | Yes | Yes | No | Yes |
| Hypokalemia | Yes | Yes | Yes | Yes |
| Hypomagnesemia | No | No | No | No |
| Hyperaldosteronism | Yes | Yes | ND | Yes |
| Hypocalciuria, NV Ca/Cr < 0.4 mg/mg | **Yes (0.01)** | **Yes (0.01)** | **Yes (0.04)** | **Yes (0.04)** |
| Renal function (eGFR) | Normal (95) | Normal (90) | Normal (92) | Normal (>90) |
| Nephrocalcinosis | No | No # | No | No |
| Neurological development | Normal | Normal | Normal | Normal |
| Sensorineural hearing loss (age at diagnosis) | **Yes (7 – 12 y)** | **Yes (2 – 6 y)** | **Yes (12 – 16 y)** | No |
| **Current treatment** | Potassium, potassium citrate, indomethacin | Potassium, potassium citrate, celecoxib | Potassium chloride, indomethacin, eplerenone | Potassium chloride, potassium citrate, indomethacin |
| **KCNJ16 variants** |  |  |  |  |
| Zygosity | Homo |  | Comp-het | Homo |
| Variant type (location) | Missense (pore) |  | Missense (pore) + nonsense (C-ter) | Nonsense (N-ter) + nonsense (N-ter) |
| Nucleotide level | c.409C>T |  | c.[395T>G]; [526C>T] | c. [142A>T; 171T>G] |
| Protein level | p.Arg137Cys |  | p.[Ile132Arg]; [Arg176*] | p.[Lys48*;Tyr57*] |

SD: standard deviation, eGFR: estimated glomerular filtration rate. This value was obtained according to the Schwartz formula in children over 1 year of age, and by CKD-EPI (Chronic Kidney Disease Epidemiology Collaboration) in adults. Normal value was considered when eGFR > 90 ml/min/1.73m<sup>2</sup>. TTKG: transtubular potassium concentration gradient. Na: sodium. K: Potassium, Cl: Chloride, Cr: creatinine, Ca: calcium. Mg: magnesium. HCO<sub>3</sub>: bicarbonate, FE: fractional excretion. NV: normal value. ND: not determined. #hyperechogenic kidneys.

We first studied two siblings (family A: patients A-II-1 and A-II-2 in Figure 1) with second-degree consanguinity. The index patient (A-II-1) was born before 40 weeks of gestation with normal weight and size. He presented between 5 - 9 months of life with vomiting and feeding difficulties. Between 15 - 19 months he consulted in the Emergency Department for fever, polyuria (14 ml/kg/hour) and polydipsia. At that time, growth failure and developmental delay were noted. Laboratory analyses revealed profound hypokalemia, mild hyponatremia and hypochloremia, compensated metabolic acidosis, elevated creatine kinase levels, and high plasma renin and aldosterone levels. The metabolic acidosis was corrected after fluid supplementation. The patient was placed on continuous oral potassium supplementation and oral indomethacin was started due to persistent polyuria. Initial electrocardiogram showed depression of the ST segment, which normalized after electrolyte correction. During follow-up persistent metabolic acidosis was noted, and potassium citrate was added to the treatment. Renal ultrasound showed normal sized kidneys without nephrocalcinosis. Between 7 – 11 years of age mild sensorineural hearing loss was detected on audiogram. Currently, he is <15 years of age and laboratory findings reveal hypocalciuria with normal renal function.

Patient A-II-2 was born at full term with normal weight and size, and no perinatal anomalies were found. Between 5 – 9 months of age the patient was admitted to the hospital with vomiting and acute bronchitis. At that time, he exhibited failure to thrive. Laboratory findings included severe hypokalemia with mild hyponatremia, hypochloremia and low serum bicarbonate levels. As in his brother, he was placed on continuous potassium supplements with potassium chloride, and due to polyuria, indomethacin was administered. Similarly, during the clinical course, metabolic acidosis was noted, and potassium citrate was added to the treatment. Indomethacin was switched to celecoxib due to a gastrointestinal hemorrhage. He is currently <10 years old and shows normal growth, psychomotor development, and renal function. However, bilateral sensorineural hearing loss was detected between 2 – 6 years of age on audiogram.

Patient B-II-1 comes from a non-consanguineous family. Between 12 – 16 years of age, he was diagnosed with bilateral neurosensorial hearing loss. Hypokalemia was detected between 22 – 26 years of age during a workup for chronic asthenia, weakness and cramps. Additional laboratory findings included metabolic acidosis and hypocalciuria. Bicarbonate and potassium oral supplements were started. During follow-up, acidosis improved, and no further bicarbonate supplementation was needed. Currently he is between 37 – 41 years old and shows normal renal function.

Patient C-II-1 is a female child of a consanguineous union. Between 10 – 14 months she presented with growth failure and weakness. Laboratory findings showed mild acute renal dysfunction with severe hypokalemia due to renal loss of potassium, mild metabolic acidosis, hypocalciuria and polyuria. The electrocardiogram on admission showed a T wave depression, which normalized after electrolyte correction. Long-term oral potassium supplements (potassium chloride and potassium citrate) and indomethacin were started. The patient’s renal function recovered after fluid and electrolyte reposition and remained normal throughout follow-up. However, the acidosis worsened during follow-up, currently requiring high doses of oral bicarbonate (up to 5 mEq/kg/day). At present, she is <5 years old and has normal growth and psychomotor development. In this patient hearing was monitored, and the most recent audiogram performed was normal. Both parents (C-I-1 and C-I-2) and her four siblings (C-II-2, C-II-3, C-II-4, C-II-5) were asymptomatic. Plasma and urine biochemical analyses were performed in all of them and yielded normal results.

### Genetic Analysis

Genetic analysis revealed *KCNJ16* pathogenic or likely pathogenic variants in all affected individuals (Figure 1 and Table. 1). In family A, a missense variant (c.409C>T, p.Arg137Cys) in the pore forming region of *KCNJ16* was identified in homozygous state in both siblings. The analysis of the parents’ DNA confirmed that the variant was present in heterozygous state in both parents. In individual B-II-1 we identified two variants: a missense variant (c.395T>G; p.Ile132Arg) in the pore region and a premature stop codon (c.526C>T;p.Arg176*) in the c-terminus of *KCNJ16*. In his mother, the missense (c.395T>G; p.Ile132Arg) variant was found in the heterozygous state, supporting the conclusion that the patient carried the variants in compound heterozygosity. All the above three variants have been previously described in other patients and are considered pathogenic [1]. In patient C-II-1, we identified two premature stop codons in the homozygous state (c.[142A>T; 171T>G], p.[Lys48*;Tyr57*]). Both the parents of this patient and the four siblings were heterozygous carriers of both variants (c.[142A>T;171T>G], p.[Lys48*;Tyr57*]). The variants are located on the same allele, as they appear in the same read in the NGS analysis. The variant p.Lys48* has been described previously and is considered pathogenic [11]. The other variant found in this patient, p.Tyr57*, has not been previously reported and *in silico* analyses predicted it to be pathogenic. Based on this information, the ACMG pathogenicity prediction tools classified the mutation p.Tyr57* as likely pathogenic, when found individually [24].

### Functional consequence of missense variants

Here, we report *KCNJ16* variants R137C, I132R, R176*, K48* and Y57*. We previously delineated the functional consequences of R137C, I132R, and R176* in Schlingmann et al., [1] In this study, we performed the functional studies for both the nonsense variants and compared the effect of nonsense variants with that of the above-mentioned variants (pore domain and c-terminal truncation mutations) of *KCNJ16* on the resulting K_ir_ channel currents.

For functional analyses of both the nonsense variants of *KCNJ16*: K48* and Y57*, the mutations were introduced into full-length cDNA encoding human *KCNJ16* and co-expressed with *KCNJ10* and *KCNJ15* in *Xenopus* oocytes. Currents were measured by two-electrode voltage clamp as described [1]. As reported previously, co-injecting cRNAs of *KCNJ16* with *KCNJ10* resulted in a strong increase in current amplitudes evoked by K_ir_4.1/K_ir_5.1 heteromers. Furthermore, heteromers of K_ir_4.1 and K_ir_5.1 (K10/K16) yielded strong inward rectifying current-voltage relationships, compared to homomers of K_ir_4.1(K10) alone (Fig. 2 A). Next, co-injecting *KCNJ10* with the nonsense variants of *KCNJ16* (K48* and Y57*) resulted in currents comparable to those elicited by K10 homomers (Fig. 2 A, B). Next, we co-injected *KCNJ16* with *KCNJ15*. As reported previously we observed a very strong increase in current amplitudes evoked by the K_ir_4.2/K_ir_5.1 (K15/K16) heteromers. Co-injecting *KCNJ15* with the nonsense variants of *KCNJ16* (K48* and Y57*) resulted in currents comparable to those elicited by K15 homomers (Fig. 2 D, E). As speculated by Webb et al., the mRNA transcripts of *KCNJ16* variants K48* and Y57* carrying premature stop are likely to result in nonsense mediated decay (NMD) in the *Xenopus* oocytes, thus promoting the homomer assembly of K10 and K15 [11, 25].

**Fig. 2.**
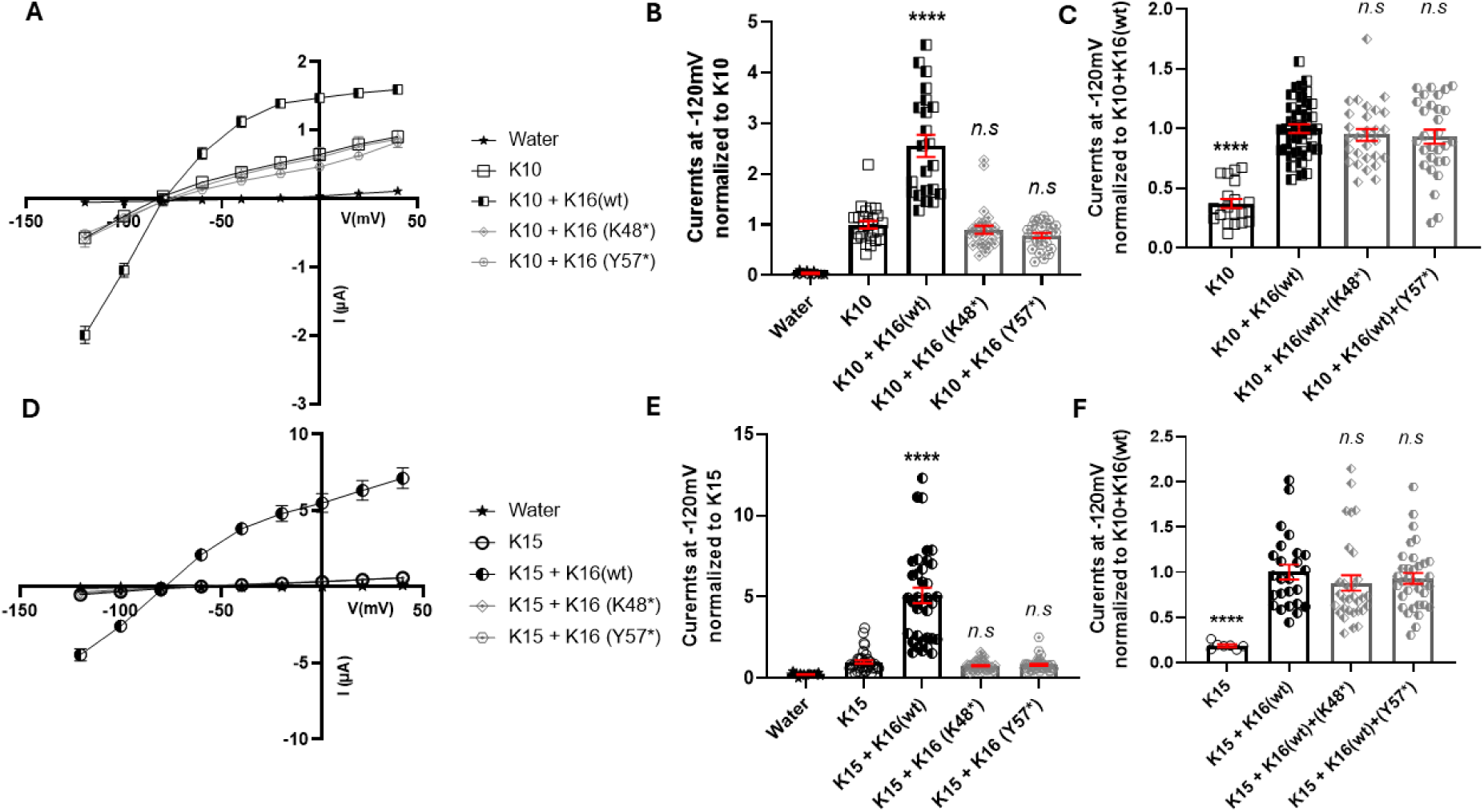
Functional consequences of *KCNJ16* (K_ir_5.1) nonsense mutations on *KCNJ10* (K_ir_4.1) and *KCNJ15* (K_ir_4.2). *KCNJ16* (K16) nonsense variants were co-expressed with *KCNJ10* (K10) or *KCNJ15* (K15) in Xenopus oocytes, either individually or in the parental allele combinations seen in family C. Oocytes were injected with K10 (1ng), K15 (2.5ng) or equimolar cRNA mixtures with WT or mutant K16 and investigated by two-electrode voltage clamp after 48 h in 2 mM bath [K^+^]. Currents were evoked in 200-ms pulses applied in 20-mV increments of potential from −120 to +40 mV (holding: −80 mV). Current-voltage curves for **(A, D)** I-V curves show inward rectification and current enhancement with WT K16. **(B, E)** Normalized currents at −120 mV and **(C, F)** Normalized currents for parental combination. Nonsense variants did not enhance K10 or K15 currents, resembling homomeric channels, whereas parental combinations resembled WT K10/K16 or K15/K16 heteromers, consistent with parents’ asymptomatic phenotype. Water-injected oocytes served as controls. Error bars represent SEM (≥10 oocytes, ≥3 batches). ****P<0.0001 and *n.s* indicate non-significant differences (*p* > 0.05).

To recapitulate the heterozygous genotype in the asymptomatic parents and other siblings from family C, we co-expressed combination of *KCNJ16* (wt and nonsense variant) together with *KCNJ10* (Fig. 2 C) and *KCNJ15* (Fig. 2 F). Here we studied both the nonsense variants K48* and Y57*. In both cases when *KCNJ16* nonsense variants (0.5ng/oocyte) were combined with *KCNJ16* wt (0.5ng/oocyte) and co-injected with *KCNJ10* (1ng/oocyte) or *KCNJ15* (1ng/oocyte), the resulting K_ir_ channel current amplitudes raised to the levels of the wild type heteromeric channels, suggesting that half an allelic copy of *KCNJ16* is necessary and enough for the constitution of heteromeric functional channels. This further rule out the presence of truncated proteins that interfere with the heteromeric assembly of the channels.

### Cellular localization of homomers of K_ir_4.1, K_ir_4.2 and K_ir_5.1 (wt and variants)

To characterize the subcellular localization of K_ir_4.1, K_ir_4.2, and K_ir_5.1, we expressed aforementioned Kir channels tagged with GFP in HeLa cells. Localization was examined 48h post-transfection using confocal microscopy. The analyses included both single transfections and co-transfections with RFP-tagged organelle markers specific for plasma membrane (PM) and endoplasmic reticulum (ER). LysoTracker dye was used to stain the lysosomes (Lyso). Distinct localization profiles were observed for each protein (Fig. 3 A, B, C). K_ir_4.1 (K10) predominantly localized to PM, exhibiting substantial co-localization with the PM marker (PLCδ-PH) (Fig. 3A1). Although partial co-localization with the ER was detected (Fig. 3 A2), there was no significant overlap with lysosomes (Fig. 3 A3). K_ir_4.2 (K15) demonstrated a similar pattern, yet less pronounced staining in the membrane as observed by a weak PM localization (Fig. 3B1) and partial co-localization with the ER marker (Fig. 3 B2) but lacked detectable lysosomal localization (Fig. 3 B3). By contrast, K_ir_5.1 (K16) showed predominant co-localization with the ER marker (Fig. 3 C2) and almost no association with the PM marker (Fig. 3 C1), indicating that K_ir_5.1 is largely retained within the ER. Moreover, K_ir_5.1 exhibited no appreciable co-localization with the lysosomal marker (Fig. 3 C3), further supporting its ER retention rather than trafficking to lysosomes.

**Fig. 3.**
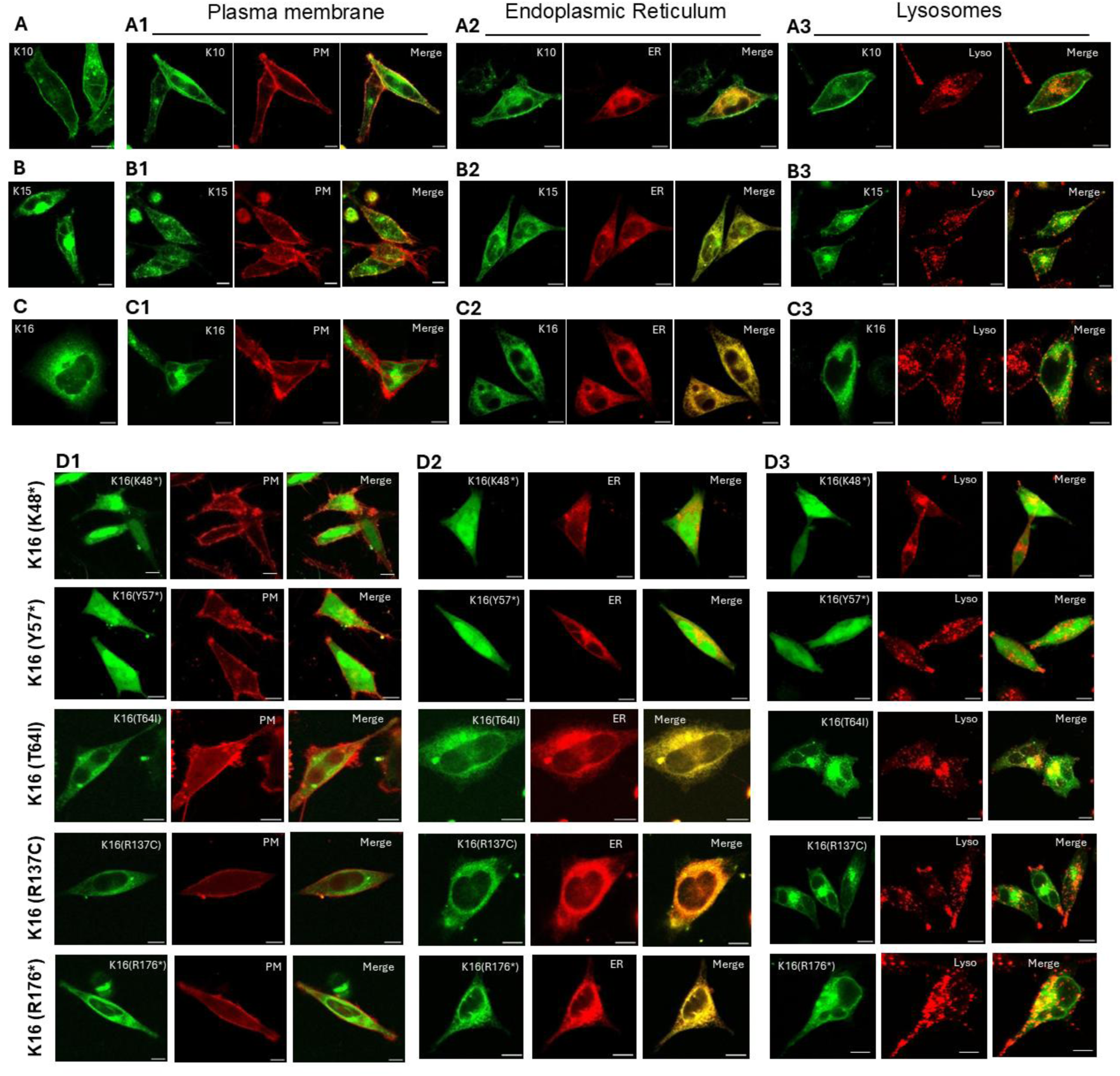
Cellular localizations of K10, K15, K16 and K16 variants. HeLa cells were transfected with GFP-tagged wild-type K10, K15, K16 or K16 variants. Panels A-C show representative images of cells expressing WT K10, K15, or K16; Panel D shows cells expressing K16 variants. For co-transfection, GFP-tagged constructs were co-expressed with PLCδPH-pRFPC3 (PM; **A1-D1)** and pDsRed2-ER (ER; **A2-D2)**. Lysosomes were labeled with LysoTracker Red DND-99 **(A3-D3)**. Merged images display co-localization in yellow. K16 variants analyzed: K16(K48*), K16(Y57*), K16(T64I), K16(R137C), and K16(R176*). Images were acquired 48 h post-transfection. Scale bar: 10 µm.

Following our findings that wild-type K_ir_5.1(K16) primarily localizes to the ER, we further investigated the subcellular localization of disease-causing K16 variants to determine if the mutations altered its cellular distribution owing to misfolding of the mutated proteins leading to subsequent degradation pathways. Included in this study are the representative N-terminus, pore region and C-terminus mutations identified in the patients re-reported here (K48*, R137C, R176*), along with the novel nonsense mutation Y57*. We also included the N-terminal missense mutation (T64I) reported previously [1]. We transiently expressed GFP-tagged *KCNJ*16 mutants – K16(K48*), K16(Y57*), K16(T64I), K16(R137C), and K16(R176*) – in HeLa cells. Their localization was visualized after 48 hours using confocal microscopy. RFP-tagged subcellular markers were used to visualize PM, ER and lysosomes. As shown in Figure 3 D1, none of the tested K_ir_5.1 (K16) mutants exhibited significant co-localization with the PM marker (PLCδ-PH). This absence of substantial PM localization for any of the mutants is consistent with the primary ER localization observed for wild-type K_ir_5.1. Furthermore, all five mutants showed predominant co-localization with the ER marker (Fig. 3 D2). The merge images clearly illustrate that the GFP signal of each mutant largely overlapped with the red fluorescence of the ER marker, indicating their primary residence within the endoplasmic reticulum. This pattern is notably similar to the established ER localization of wild-type K_ir_5.1. Crucially, we assessed potential trafficking to lysosomes, a common pathway for the degradation of misfolded or improperly trafficked ion channel mutants. As depicted in Figure 3 D3, none of the K_ir_5.1 mutants demonstrated substantial co-localization with the lysosomal marker. This finding is significant as it rules out the common dogma of a targeted lysosomal degradation of misfolded disease-causing mutations in ion channels [26]. In summary, the subcellular localization analysis of five different K_ir_5.1 mutants revealed that their primary localization remained within the ER, mirroring that of wild-type K_ir_5.1. None of the mutants showed significant trafficking to the PM or, notably, to lysosomes, indicating that these representative disease-causing mutations do not alter the fundamental intracellular sorting and retention mechanisms of K_ir_5.1 towards lysosomal degradation. This suggests that the pathogenic mechanisms associated with these mutations may not involve a classical misfolding-induced lysosomal degradation pathway.

### Effect of disease-causing variants in K_ir_5.1 on the membrane localization of K_ir_4.1/K_ir_5.1 and K_ir_4.2/K_ir_5.1 hetero tetramers

Building upon our previous observations regarding the primary ER localization of wild-type K_ir_5.1 (K16), we next investigated how co-expression with K_ir_4.1 (K10) and K_ir_4.2 (K15) impacts the cellular distribution of K_ir_5.1, and how disease-causing K_ir_5.1 mutations affect both heteromerization and the subcellular localization of hetero tetramers of K_ir_4.1/K_ir_5.1(K10/K16) and K_ir_4.2/K_ir_5.1(K15/K16). We observed that wt K16, when co-expressed with either K10 or K15, exhibited a striking shift from its characteristic ER retention to the plasma membrane (PM), displaying clear co-localization of K16 with K10 (Fig. 4 A1) and K15 (Fig. 4 B1) at the cell surface. This strongly suggested that the formation of heteromeric channels involving K16 and either K10 or K15 facilitated the trafficking of K16 to the PM. Of particular importance is the strong enhancement of the PM localization of heteromeric channels of K15/ K16 (Fig. 4 B1, compared with the homomers of K15 (Fig. 3 B, B1).

**Fig. 4.**
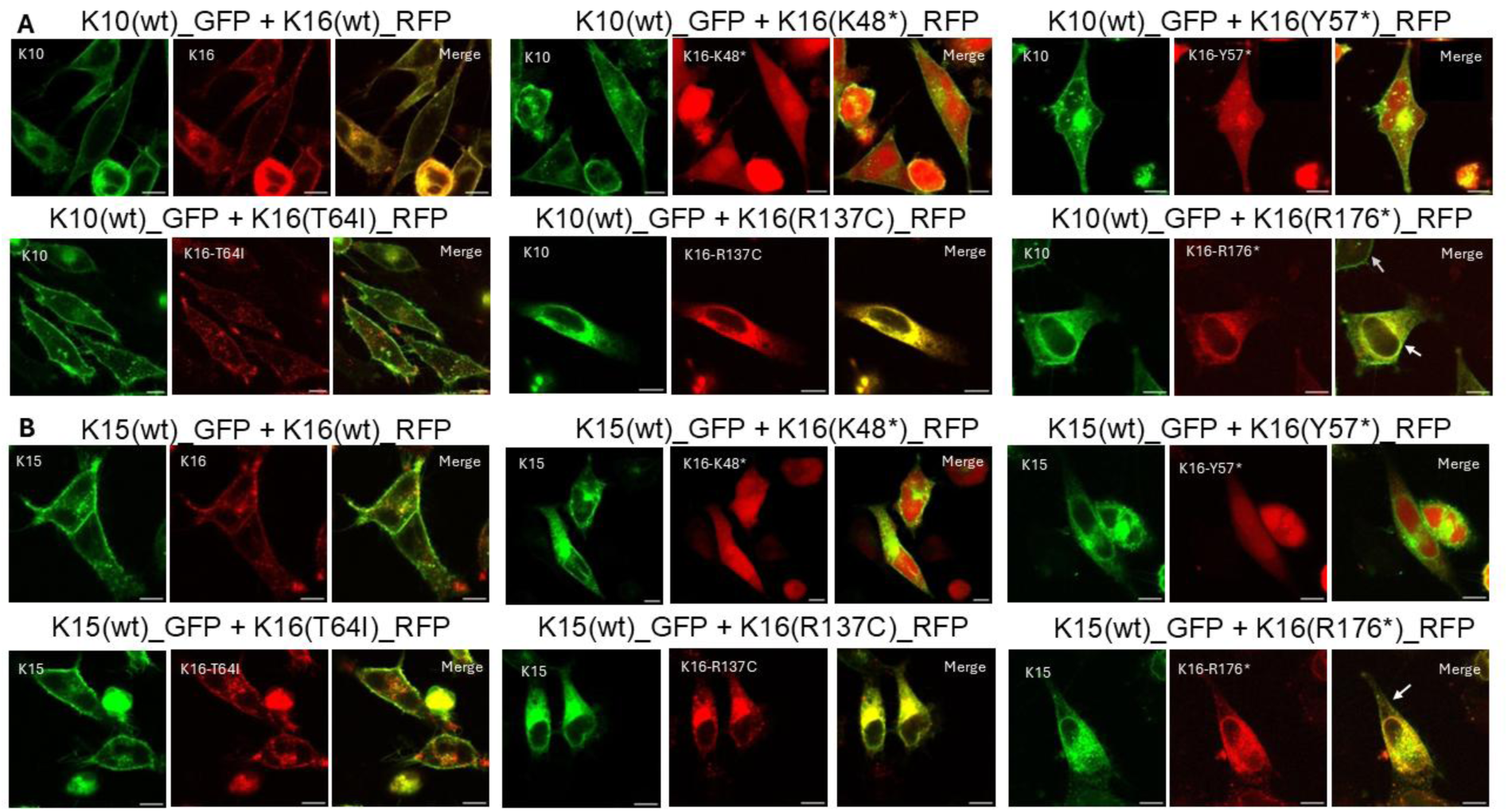
Membrane localization of K10 and K15 co-expressed with K16 WT and variants. HeLa cells were co-transfected with GFP-tagged K10 or K15 (green) and RFP-tagged K16 WT or variants (red) to assess the effect of K16 WT or variants on K10/K16 (A1-6) or K15/K16 (B1-6) heteromer localization. Merged images highlight co-localization in yellow. Images were acquired 48 h post-transfection. Scale bar: 10 µm.

We then assessed the impact of disease-causing variants of K16 on the heteromerization and cellular distribution of K10 and K15. The heteromers involving the N-terminal mutant K10/ K16(T64I) (Fig. 4 A4) or K15/ K16(T64I) (Fig. 4 B4), largely retained their ability to traffic to the PM, albeit with slightly reduced PM presence compared to wild-type heteromers (Fig. 4 A1 and B1 respectively). In stark contrast, the N-terminal early stop nonsense mutant proteins K16(K48*) and K16(Y57*) showed diffused intracellular staining (comparable to control GFP) when co-expressed with K10 (Fig. 4 A2 and A3 respectively) or K15 (Fig. 4 B2 and B3 respectively). Moreover, membrane localization of K10 and K15 resembled that of their homomeric channels, suggesting that the premature stop variants resulted in complete loss of K16 which severely compromised the enhanced PM targeting of K10 or K15. Furthermore, both the pore-lining mutant K16(R137C) and C-terminal truncated protein K16(R176*), while primarily localized to the ER similar to the base line of the wild-type K16, had strikingly different effects on K10 (Fig. 4 A5, A6) and K15 (Fig. 4 B5, B6) when compared with their wild-type counterpart. While wild-type K16 enhanced the PM targeting of K10 and K15, pore-mutant (R137C) strongly retained both K10 and K15 within intracellular compartments, specifically the ER, with no discernible PM trafficking (Fig. 4 A5 and B5 respectively). On the other hand, although the C-terminal truncation mutation (R176*) also strongly retained both K10 and K15 in the ER (Fig. 4 A6 and B6 respectively), we observed faint membrane staining of the heteromeric channels of K10/K16(R176*) and K15/K16(R176*) as shown by white markers in Figures 4 A6 and B6 respectively. Note the difference in strength of membrane localization of K10/K16(R176*) heteromers (white marking) Vs. K10 homomer (gray marking) in the figure 4 A6. Similar effect was also observed with C-terminal missense mutation K16(P250L) (See SI-Fig.1A and B).

### Effect of K_ir_5.1 variants on protein-protein interaction with K_ir_4.1 and K_ir_4.2

We performed Co-immunoprecipitation (Co-IP) experiments in HEK293 cells to determine whether disease causing mutations in K_ir_5.1(K16) affects its ability to heteromerize with K_ir_4.1 (K10) and K_ir_4.2 (K15). For Co-IP, Flag-tagged K_ir_5.1(K16-Flag) was co-expressed with either GFP-tagged K_ir_4.1(K10-GFP) or GFP-tagged K_ir_4.2 (K15-GFP). As shown in Figures 5 A1 and A3 (green arrows), K16-Flag was efficiently immunoprecipitated in both the input (IP) and co-IP fractions, confirming the effectiveness of the anti-Flag beads for pulldown. Furthermore, bands corresponding to K10-GFP (Fig. 5 A2, red arrow) and K15-GFP (Fig. 5 A4, red arrow) were detected in the co-immunoprecipitated samples, indicating that both K10-GFP and K15-GFP specifically co-immunoprecipitated with K16-Flag. These findings strongly suggest that K16 heteromerized with both K10 and K15 when co-expressed in HEK293 cells. To validate the specificity of these interactions, control experiments were performed in which either the Flag- or GFP-tagged constructs were expressed alone (blue arrows in Fig. 5 B1–4). As expected, these control conditions showed no detectable bands for potential interaction partners, confirming the absence of non-specific co-immunoprecipitation and supporting the specificity of the observed protein-protein interactions.

**Fig. 5.**
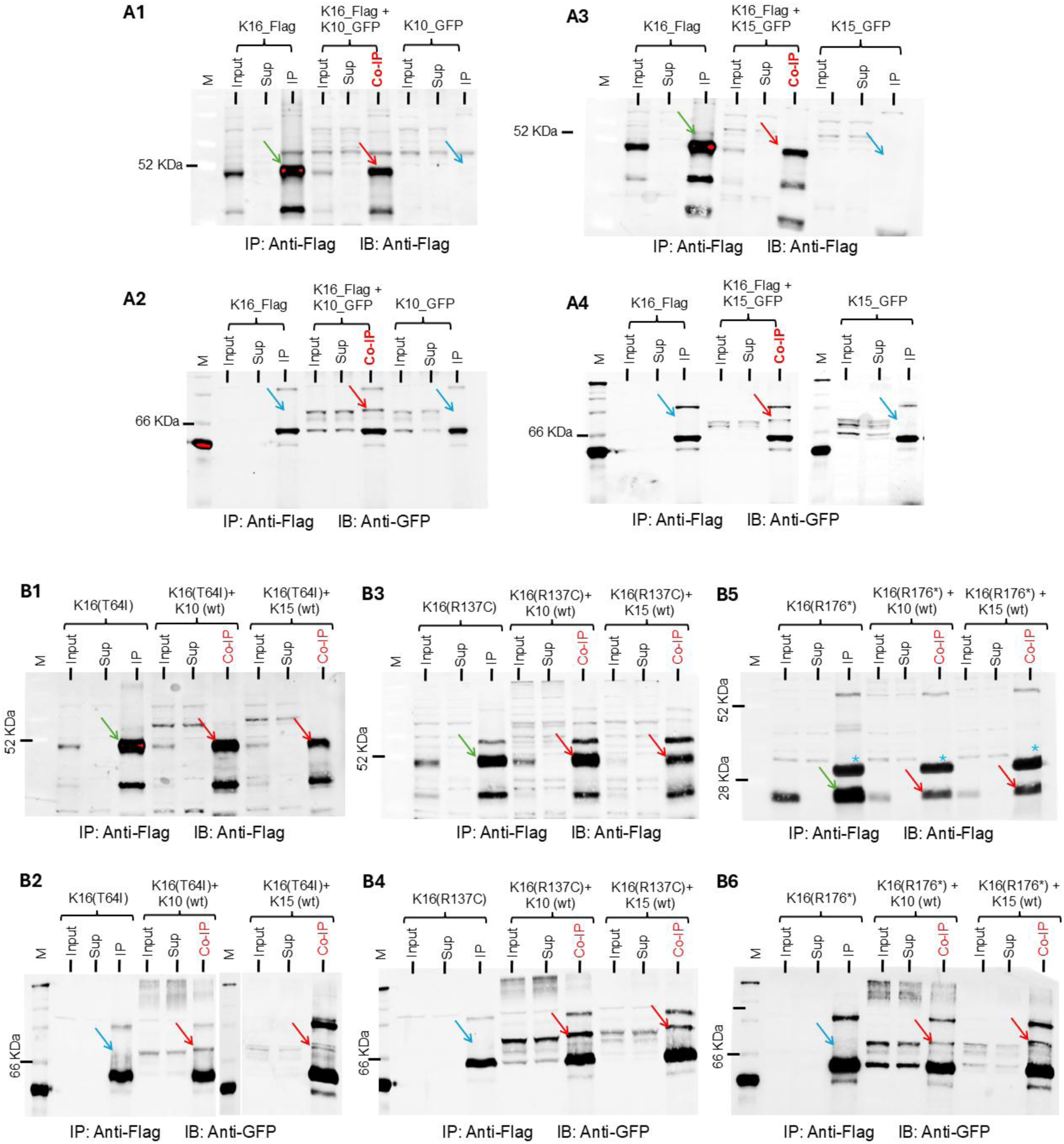
Effect of K16 variants on interaction with K10 and K15. Co-immunoprecipitation assays in HEK293 cells examined interactions between Flag-tagged K16 and GFP-tagged K10 or K15. Panels **A1-A4 show** Co-Ips with WT K16 and panels **B1-B6** show Co-Ips with K16(T64I), K16(R137C) and K16(R176*). Cell lysates were immunoprecipitated with anti-Flag beads, and western blots were probed with anti-Flag to detect K16-Flag and anti-GFP to detect co-immunoprecipitated K10-GFP or K15-GFP. No bands were detected in controls lacking Flag- or GFP-tagged proteins, confirming Co-IP specificity. Expected molecular weights: ∼50 kDa (K16-Flag WT, T64I, R137C), ∼28 kDa (R176*) and ∼70 kDa (K10-GFP, K15-GFP).

Having established that wild-type K16 heteromerized with K10 and K15, we next investigated whether disease-causing variants of K16 retain this ability. To address this question, we conducted Co-IP assays in a manner similar to that described above but expressing K16 variants (T64I, R137C, and R176*). We used the wild-type K16 as a control. As shown in Figures 5 B1, B3, and B5, all Flag-tagged K16 constructs (wt and variants), were efficiently immunoprecipitated via anti-Flag affinity purification (green and red arrows in the IP and Co-IP fractions respectively). This demonstrates that the bait proteins were recovered effectively under the assay conditions. Importantly, in co-expression experiments, strong bands corresponding to co-immunoprecipitated K10-GFP and K15-GFP were observed in the Co-IP fractions for all tested K16-flag variants (red arrows in Fig. 5 B2, B4, and B6). Specifically, K10-GFP co-immunoprecipitated robustly with K16(T64I)-Flag (Fig. 5 B2), K16(R137C)-Flag (Fig. 5 B4), and K16(R176*)-Flag (Fig. 5 B6). Similarly, K15-GFP also co-immunoprecipitated with all three Flag-tagged K16 variants (Fig. 5 B2, B4, B6). These results indicated that despite the presence of point mutations (T64I and R137C) or truncation (R176*), all tested K16 variants maintained their ability to heteromerize with both K10 and K15, when co-expressed in HEK293 cells. Similar to the wild-type experiments, control conditions in which only one of the tagged proteins was expressed (blue arrows in Fig. 5 B2, B4 and B6) showed no evidence of non-specific binding, further confirming the specificity of the observed protein complex formation. Additionally, Co-IP experiments performed with the C-terminal missense variant K16(P250L) showed similar results, suggesting interaction of K16(P250L)-Flag with K10-GFP and K15-GFP (See SI Fig. 1 C, D).

## Discussion

Inward rectifying potassium (K_ir_) channels are found in almost every cell type in the human body and are indispensable in regulating electrolyte homeostasis, membrane potential and cell excitability [27]. Especially in the kidney they are found in both apical and basolateral compartments of polarized tubular epithelia and aid in potassium recycling necessary for salt reabsorption [1]. Pathogenic variants of these K_ir_ channels are associated with renal salt wasting disorders such as Bartter type-II (K_ir_1.1 encoded by *KCNJ1*) and EAST/SeSAME (K_ir_4.1 encoded by *KCNJ10*) syndromes [18]. Both disorders feature hypokalemia usually indicating a compensatory activation of RAAS leading to metabolic alkalosis. In EAST/SeSAME syndrome, mutations in *KCNJ10* affect the function of both homomeric K_ir_4.1 as well as heteromeric K_ir_4.1/ K_ir_5.1channels, which comprise the major basolateral potassium channels in DCT [9, 10]. Recently, disease-causing mutations in *KCNJ16* have been added to the growing array of renal tubular salt wasting disorders [1, 11]. Patients with mutations in *KCNJ16* develop a complex tubulopathy characterized by hypokalemia, salt wasting, varying severity of acidosis and sensorineural deafness, sometimes referred to as HkTD (Hypo kalemic Tubulopathy and Deafness) [1, 11, 13]. Interestingly, symptomatology of these patients does not coincide with that of EAST/SeSAME syndrome. Furthermore, the renal phenotype of both HkTD patients and *KCNJ16*^-/-^ animal is somewhat peculiar with an unusual combination of hypokalemia and acidosis. Unlike the kidney, expression of K_ir_5.1 in the brain does not completely overlap with that of K_ir_4.1 [2, 27]. This explains the lack of Epilepsy and Ataxia in HkTD patients. Moreover, expression of K_ir_5.1 overlaps that of K_ir_4.2 in the proximal tubule [8]. Functional studies by Schlingmann et al., showed that besides their effect on K_ir_4.1, variants of K_ir_5.1 decrease the channel activity of K_ir_4.2/ K_ir_5.1 heteromers as well [1]. This is reminiscent with the genetic ablation studies in mice exhibiting hyperchloremic metabolic acidosis, reduced threshold for bicarbonate resorption and decreased urinary ammonia excretion [8], pointing towards extra-distal hypokalemia and proximal tubular acidosis.

Several groups have reported pathogenic to benign mutations in *KCNJ16* gene (Table. 2). Till date 13 mutations in *KCNJ16* have been reported. Of which, six variants (in diverse zygosities) were reported in HkTD patients with varying degrees of acidosis, one variant in a HkTD patient with alkalosis, and one benign variant and two variants reported in SIDS patients [1, 11, 13, 14, 17, 28]. In the present study, we report 4 patients from 3 families of both consanguineous and non-consanguineous unions. At the time when first symptoms were observed their ages ranged from 7m to 25y and displayed renal phenotype featuring salt wasting, hypokalemia and varying degrees of acidosis. Here, we re-report two gene variations in *KCNJ16* which we formerly described in Schlingmann et al.These variants are located in the so-called mutational hotspot regions of the K_ir_ family (along the G-loop and adjacent C-terminal regions) [18]. The clinical symptoms of the patient carrying the homozygous R137C variant coincided with our previously reported ones. However, the clinical phenotype of the patient carrying the compound heterozygous I132R+R176* variants showed stark deviation from the ones we reported previously. The phenotype in this patient not only developed later in age (over 20 y of age) but also showed severe yet transient acute acidosis, that required no further bicarbonate supplementation. Furthermore, the patient exhibited normal growth (+0.8 SD), contrasting with the short stature and failure to thrive typically observed in patients with renal tubular acidosis [29], thus supporting the late-onset and transient course of acidosis in this case. This further adds to the granular clinical picture of HkTD (See Table. 2). Here we also report a novel biallelic nonsense mutation (Y57*), which further expands the variant spectrum of *KCNJ16*. This patient C-II-1 featured two nonsense mutations at positions 48 (previously reported by Webb et al.,) and 57 in homozygous state, resulting in complete loss of K_ir_5.1 channels. Importantly, the patient showed strong hypokalemia and only mild acidosis initially but developed severe phenotype during the follow-up.

**Table. 2.**
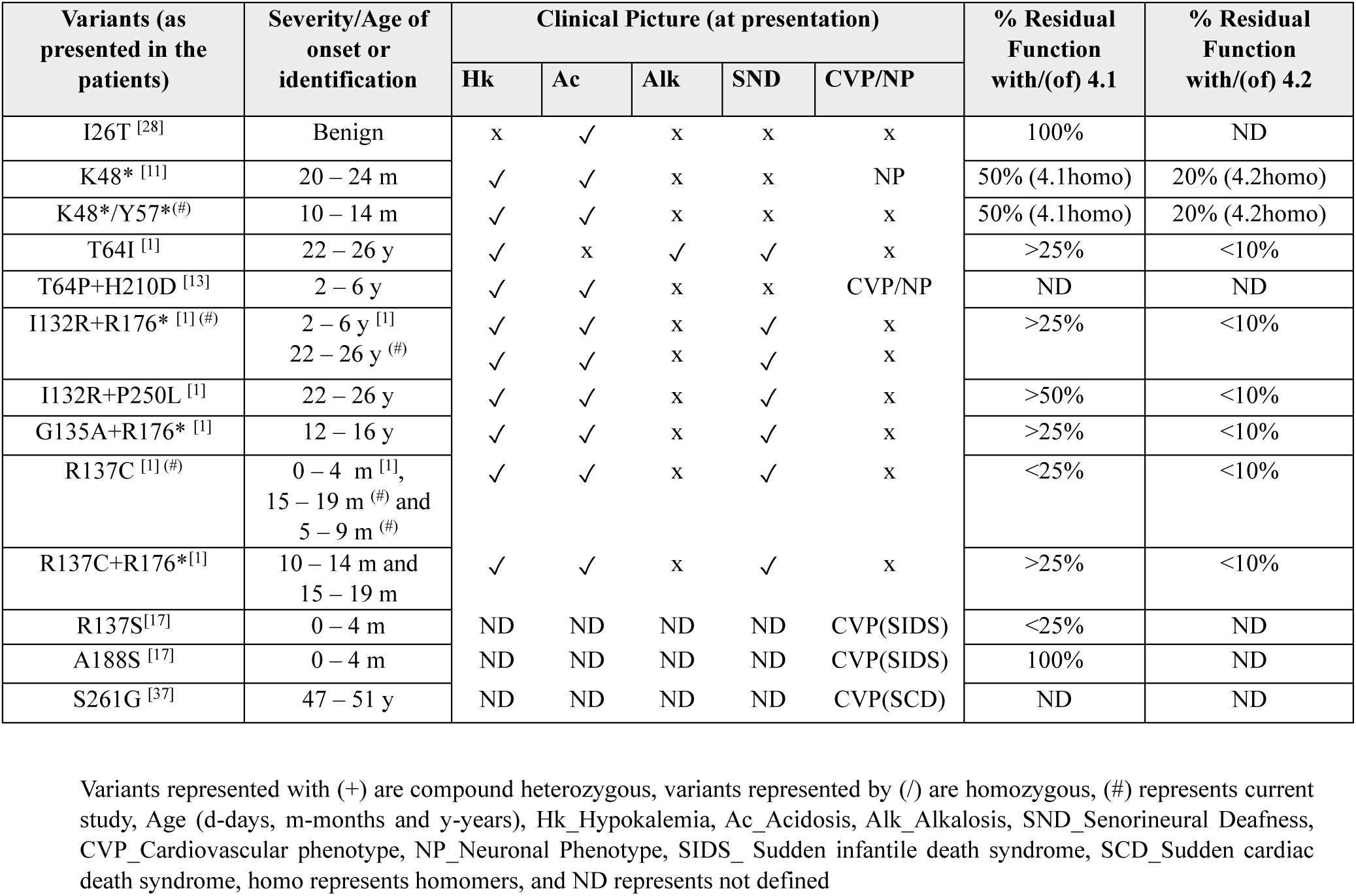
*KCNJ16* variants and their consequences.

Here, functional studies were performed for both nonsense variants (K48* and Y57*) and compared the effects with that of the other variants observed in patients from families A (R137C) and B (I132R/R176*). Our data showed that indeed, both nonsense variants of K_ir_5.1 (K16), resulted in currents similar to K_ir_4.1 (K10) or K_ir_4.2 (K15) homomers. Therefore, dominant negative influence of small, truncated proteins on the homomeric assembly of K10 or K15 can be ruled out in favor of the prevalence of nonsense mediated decay of the transcript RNA carrying premature termination. Hence, one would expect a milder phenotype in these patients with N-ter nonsense mutations, owing to about 50% channel function of K10 homomers and about 10% channel function of K15 homomers, in the absence of K16. However, this is not reflected in the clinical picture of patient C-II-1. Therefore, the severe hypokalemia and acidosis observed in this patient (in line with Webb et al.,) underscores not only the essential role of Kir5.1 in renal transport mechanisms but also suggests the involvement of additional, as yet unidentified, Kir5.1-related pathways. We previously showed that variants R137C and I132R+R176*, presented with severe hypokalemia and acidosis. Both our patients (A-II-1 and A-II-2) with variant R137C are reminiscent of the previously reported where we showed that R137C resulted in a marked reduction (<25% residual K_ir_ channel activity) in the resulting heteromeric K10/K16(R137C) currents when compared to that of the K10 homomers (Table. 2). However, compound heterozygotes expressing both I132R and R176* heteromerized with K10 yielded currents that were only slightly lower than that of K10 homomers (Table. 2). This divergent phenotype of the patient B-II-1 compared with the two patients previously reported, once again suggests that the importance of K16 in renal transport mechanisms is not understood to its actual completeness. Lastly, K16 variants had a much stronger effect on K15 than on K10, which explains the strong acidosis seen in these patients despite hypokalemia [1].

In addition to hypokalemia and varying severity of acidosis, patients with *KCNJ16* variants also featured in common sensorineural deafness. K_ir_4.1 is expressed in the intermediate cells of the stria vascularis and is responsible for maintaining high [K^+^] in the endolymph and thereby maintaining the endocochlear potential [27, 30]. Mutations in K_ir_4.1 leads to sensorineural deafness in EAST/SeSAME patients [9, 10]. The expression pattern of Kir5.1 does not coincide with that of K_ir_4.1 in the inner ear. On the other hand, K_ir_5.1 is expressed in the root cells and spindle cells of the spiral ligament where its expression coincides with that of K_ir_4.2 [27, 31]. Lack of disease-causing mutations in K_ir_4.2 limits our knowledge about its role in auditory system in humans. Furthermore PSD-95, that regulates Kir5.1 membrane trafficking and function is also expressed in the inner and outer hair cells [5, 32]. However, mice lacking *KCNJ16* reflected that Kir5.1 is nonessential for auditory functions in mice. Hence, unless the exact role of K_ir_5.1 in the sensory transduction of hearing is not clearly elucidated, the occurrence of sensorineural deafness observed in most HkTD patients remains poorly understood. On these lines, it is important to highlight that the patient C-II-1, did not show signs of sensorineural deafness (till date) unlike other patients of similar age in our cohort and elsewhere reported [1, 11]. Furthermore, Webb et al., have also not mentioned hearing impairment in their patient affected by the biallelic loss of function variant (K48*). This is in line with the *Kcnj16^-/-^* mouse where absolute lack of K_ir_5.1 did not alter their auditory function [33]. Put together nonsense mutations in *KCNJ16* resulting in complete absence of K_ir_5.1 may have little effect on sensorineural hearing in these patients owing to residual function of the interacting K_ir_ channel involved in the auditory system. On the other hand, missense mutations may have a dominant negative effect on the same resulting in deafness.

Considering the widespread distribution of K_ir_5.1 channels which coincides with that of the interacting heteromer partners and the fact that although K16 can form homomers, their membrane integration requires heteromerization or association with modulator proteins, it is tempting to speculate that the variability of the clinical picture is embedded in the nature of protein-protein interaction and membrane localization of functional channels. To explore this, we performed Co-immunoprecipitation studies in HEK293 cells to study protein-protein interactions between K_ir_5.1 (K16) (wt and mutant) and its heteromeric partners K_ir_4.1 (K10) and K_ir_4.2 (K15). All variants including the C-terminal truncated R176* interacted with both K10 and K15. This is consistent with the established understanding that heteromeric assembly of Kir channels is not limited to a single domain but extends across multiple regions [34]. This absence of loss of interaction argues for a dominant negative effect of the K16 variants on K10 and K15. To further investigate the extent of dominant negative effect caused by K16 variants on the interacting Kir, we further investigated the cellular localization of florescence labeled homomeric and heteromeric channels of K16 (wt and mutants), K10 and K15 in HeLa cells using live cell confocal imaging. Selected variants for this study are representatives of mutation hotspots: pore region (R137C of family A) and the adjacent C-terminus (R176* of family B). Included are the nonsense mutants from Family C. Furthermore, we have also included the N-terminal variant T64I presenting with alkalosis instead of acidosis and the P250L variant (associated with milder phenotype and pathophysiology) as a representative of a C-terminal missense variant (juxtaposing the nonsense variant R176*). Subcellular localization of K10, K15 and K16 revealed distinct localization profile for each homomer. K10 homomers predominantly localized to the PM, K16 homomers showed weak PM localization and K16 homomers were majorly localized in the ER and not the lysosomes, suggesting an ER retention that accounts to their electrically silent nature, rather than lysosomal degradation. Additionally, variants of K16 also primarily localized in the ER, mirroring that of wild-type K16 homomers. We observed no colocalization of the mutant variants of K16 with lysosomes. This finding is of significance because, while disease-causing mutations in ion channels often lead to misfolding and subsequent retention in the ER [26], they can also be targeted for lysosomal degradation via pathways like ER-associated degradation (ERAD) or autophagy upon failed ER quality control [26]. The consistent absence of lysosomal co-localization for these K16 mutants suggests that their ER retention is not primarily resolved by lysosomal targeting for degradation. Instead, they remain localized within the ER, like the wild-type protein, potentially due to a yet to be explained mechanism of retention or alternative clearance pathways that do not involve bulk lysosomal degradation. Furthermore, upon co-expression with K10 or K15, K16 exhibited a striking shift from its characteristic ER retention to the PM. This strongly suggested that heteromerization of K16 with either K10 or K15 facilitates the trafficking of K16 to PM, potentially by masking an intracellular retention signal on K16 or by inducing conformational or stability changes that enable its transport as part of a hetero-tetrameric complex. Of particular importance is the strong enhancement of the PM localization of heteromeric channels K15/K16, unlike the weak PM localization of K15 homomers. Furthermore, we observed differential membrane translocation of heteromeric channels involving disease causing variants of K16. While co-expression with nonsense variants of K16 showed preservation of membrane localization of K10 and K15 homomers, co-expression with K16(R137C) resulted in their strong retention in ER. However, when co-expressed with T64I we observed robust membrane localization of K10 and K15. Also, co-expression with R176* and P250L resulted in a modest membrane localization of K10 and K15. Our data strongly suggested that the ER retention of the heteromeric channels involving mutant K16 are variant dependent. While heteromeric channels involving pore variant of K16 were stringently retained in the ER, those involving N and C-terminal variants, showed varying degrees of membrane translocation of the heteromeric channels. Thus, our study points towards a varying degree of dominant negative effect of K16 variants on the membrane translocation of the resulting heteromeric Kir channels, which in turn reflects in the clinical phenotypes of the variants (Table. 2).

There is currently no data on the long-term kidney function of patients with HkTD. In our cohort, all patients had normal renal function even after 10 years of evolution. The lack of involvement of the Henle’s loop is expected to be associated with a milder degree of polyuria and, consequently milder degree of hyperaldosteronism without nephrocalcinosis which can be related to a better prognosis of renal function (as it also occurs in patients with Gitelman syndrome [35]). Long-term follow-up studies are required to better understand renal prognosis in HkTD, although genotype-phenotype correlations regarding long-term outcome are often difficult to establish in hereditary salt-losing tubulopathies [36] due to their low prevalence throughout different populations.

## Conclusion

In summary, we report two already described variants of *KCNJ16*, with one consistent (R137C) and one divergent phenotype (I132R+R176*). Furthermore, we contribute to the expanding variant spectrum of *KCNJ16* by reporting a new nonsense variant (Y57*) lacking deafness. Alongside the functional characterization of the pathophysiology of *KCNJ16* variants highlighting the importance of K_ir_5.1 (K16) in renal health, we show that the disease-causing variants of K_ir_5.1 preserve their ability to interact with their heteromeric partners K_ir_4.1(K10) and K_ir_4.2 (K15). Yet, they have differential effects on the membrane translocation of the resulting heteromeric channels, further explaining the granular clinical picture. Finally, our study underscores the importance of elucidating the molecular mechanisms underlying *KCNJ16* variants to enable accurate diagnosis and the development of personalized therapeutic strategies. This is particularly relevant when exploring the therapeutic potential of heteromeric Kir channels and while developing interventions involving pharmacological agents such as Kir channel modulators [19].

## Data Availability

All data produced in the present study are available upon reasonable request to the authors

## Acknowledgements

The authors thank the patients and their parents for participating in this study. We thank Prof. Dominik Oliver for providing essential resources for this research. Experimental assistance from Anna Zimmermann, and Rajeshwari Bisen (Philipps University Marburg) is deeply appreciated. We thank Galina Zielke for excellent technical support. We thank the collaborating physicians of the Renaltube platform (www.renaltube.com).

## Statements of Declarations

### Author contributions

A.R, L.M designed the study. G.A, L.G.S, P.T.C, G.F.S are treating physicians. S.M, A.R, V.R, F.S, L.G, A.G.C, A.S.R, J.P.G, M.G.A, S.G.C, A.C.A.D, I.G.S carried out experiments. A.R, V.R, S.M analyzed data. A.R, S.M, I.G.S, L.G made figures. A.R, L.G wrote the original draft. A.R, V.R, L.G, L.M edited the draft. A.G.C, J.P.G, M.K, S.W, revised the paper and all authors approved the final version of the manuscript.

### Ethics approval

Ethical approval for this study was obtained from the Ethics Committee for Clinical Research of Euskadi (CEIC-E). The code for the ethics committee is PI2017118. The research was carried out in accordance with the Declaration of Helsinki on human experimentation of the World Medical Association.

### Consent to participate and publish

Before inclusion in the study, informed consent was obtained from each proband or their legal tutors and their families for both participation and publication in Fig.1.

## References

1. Schlingmann KP, Renigunta A, Hoorn EJ, Forst AL, Renigunta V, Atanasov V, et al. Defects in KCNJ16 Cause a Novel Tubulopathy with Hypokalemia, Salt Wasting, Disturbed Acid-Base Homeostasis, and Sensorineural Deafness. J Am Soc Nephrol. 2021;32(6):1498–512. doi: 10.1681/ASN.2020111587.

2. Lo J, Forst AL, Warth R, Zdebik AA. EAST/SeSAME Syndrome and Beyond: The Spectrum of Kir4.1- and Kir5.1-Associated Channelopathies. Front Physiol. 2022;13:852674. doi: 10.3389/fphys.2022.852674.

3. Starremans PG, van der Kemp AW, Knoers NV, van den Heuvel LP, Bindels RJ. Functional implications of mutations in the human renal outer medullary potassium channel (ROMK2) identified in Bartter syndrome. Pflügers Archiv - European Journal of Physiology. 2014;443(3):466–72. doi: 10.1007/s004240100708.

4. Simon DB, Karet FE, Rodriguez-Soriano J, Hamdan JH, DiPietro A, Trachtman H, et al. Genetic heterogeneity of Barter’s syndrome revealed by mutations in the K+ channel, ROMK. Nature Genetics. 1996;14(2):152–6. doi: 10.1038/ng1096-152.

5. Zhang C, Guo J. Diverse functions of the inward-rectifying potassium channel Kir5.1 and its relationship with human diseases. Front Physiol. 2023;14:1127893. doi: 10.3389/fphys.2023.1127893.

6. Pessia M, Imbrici P, D’Adamo MC, Salvatore L, Tucker SJ. Differential pH sensitivity of Kir4.1 and Kir4.2 potassium channels and their modulation by heteropolymerisation with Kir5.1. J Physiol. 2001;532(Pt 2):359–67. doi: 10.1111/j.1469-7793.2001.0359f.x.

7. Lagrutta AA, Bond CT, Xia XM, Pessia M, Tucker S, Adelman JP. Inward Rectifier Potassium Channels. Cloning, Expression and Structure-Function Studies. Japanese Heart Journal. 1996;37(5):651–60. doi: 10.1536/ihj.37.651.

8. Bignon Y, Pinelli L, Frachon N, Lahuna O, Figueres L, Houillier P, et al. Defective bicarbonate reabsorption in Kir4.2 potassium channel deficient mice impairs acid-base balance and ammonia excretion. Kidney International. 2020;97(2):304–15. doi: 10.1016/j.kint.2019.09.028.

9. Bockenhauer D, Feather S, Stanescu HC, Bandulik S, Zdebik AA, Reichold M, et al. Epilepsy, ataxia, sensorineural deafness, tubulopathy, and KCNJ10 mutations. N Engl J Med. 2009;360(19):1960–70. doi: 10.1056/NEJMoa0810276.

10. Scholl UI, Choi M, Liu T, Ramaekers VT, Häusler MG, Grimmer J, et al. Seizures, sensorineural deafness, ataxia, mental retardation, and electrolyte imbalance (SeSAME syndrome) caused by mutations in KCNJ10. Proceedings of the National Academy of Sciences. 2009;106(14):5842–7. doi: 10.1073/pnas.0901749106.

11. Webb BD, Hotchkiss H, Prasun P, Gelb BD, Satlin L. Biallelic loss-of-function variants in KCNJ16 presenting with hypokalemic metabolic acidosis. Eur J Hum Genet. 2021;29(10):1566–9. doi: 10.1038/s41431-021-00883-0.

12. Tanemoto M, Abe T, Onogawa T, Ito S. PDZ binding motif-dependent localization of K+ channel on the basolateral side in distal tubules. Am J Physiol Renal Physiol. 2004;287(6):F1148–53. doi: 10.1152/ajprenal.00203.2004.

13. Chen J, Fu Y, Sun Y, Zhou X, Wang Q, Li C, et al. Novel KCNJ16 variants identified in a Chinese patient with hypokalemic metabolic acidosis. Molecular Genetics & Genomic Medicine. 2023;11(11). doi: 10.1002/mgg3.2238.

14. Zhang J, Han J, Li L, Zhang Q, Feng Y, Jiang Y, et al. Inwardly rectifying potassium channel 5.1: Structure, function, and possible roles in diseases. Genes & Diseases. 2021;8(3):272–8. doi: 10.1016/j.gendis.2020.03.006.

15. Xu B, Dissanayake LV, Levchenko V, Zietara A, Kravtsova O, Staruschenko A. Deletion of Kcnj16 altered transcriptomic and metabolomic profiles of Dahl salt-sensitive rats. iScience. 2024;27(10):110901. doi: 10.1016/j.isci.2024.110901.

16. Poli G, Hasan S, Belia S, Cenciarini M, Tucker SJ, Imbrici P, et al. Kcnj16 (Kir5.1) Gene Ablation Causes Subfertility and Increases the Prevalence of Morphologically Abnormal Spermatozoa. Int J Mol Sci. 2021;22(11). doi: 10.3390/ijms22115972.

17. Neubauer J, Forst A-L, Warth R, Both CP, Haas C, Thomas J. Genetic variants in eleven central and peripheral chemoreceptor genes in sudden infant death syndrome. Pediatric Research. 2022;92(4):1026–33. doi: 10.1038/s41390-021-01899-4.

18. Zangerl-Plessl E-M, Qile M, Bloothooft M, Stary-Weinzinger A, van der Heyden MAG. Disease Associated Mutations in KIR Proteins Linked to Aberrant Inward Rectifier Channel Trafficking. Biomolecules. 2019;9(11). doi: 10.3390/biom9110650.

19. McClenahan SJ, Kent CN, Kharade SV, Isaeva E, Williams JC, Han C, et al. VU6036720: The First Potent and Selective In Vitro Inhibitor of Heteromeric Kir4.1/5.1 Inward Rectifier Potassium Channels. Mol Pharmacol. 2022;101(5):357–70. doi: 10.1124/molpharm.121.000464.

20. Shaikh IG, Kostritskii AY, Renigunta A, Jeschke J, Halaszovich CR, Zhao W, et al. Electrochemical coupling at the plasma membrane by mouse voltage-sensitive phosphatase requires association with basigin. Cell Rep. 2025;44(9):116200. doi: 10.1016/j.celrep.2025.116200.

21. Shaikh IG, Kostritskii AY, Renigunta A, Jeschke J, Halaszovich CR, Zhao W, et al. Electrochemical coupling at the plasma membrane by mouse voltage-sensitive phosphatase requires association with basigin. Cell Reports. 2025;44(9). doi: 10.1016/j.celrep.2025.116200.

22. Renigunta V, Xhaferri N, Shaikh IG, Schlegel J, Bisen R, Sanvido I, et al. A versatile functional interaction between electrically silent KV subunits and KV7 potassium channels. Cellular and Molecular Life Sciences. 2024;81(1). doi: 10.1007/s00018-024-05312-1.

23. Renigunta A, Renigunta V, Saritas T, Decher N, Mutig K, Waldegger S. Tamm-Horsfall Glycoprotein Interacts with Renal Outer Medullary Potassium Channel ROMK2 and Regulates Its Function. Journal of Biological Chemistry. 2011;286(3):2224–35. doi: 10.1074/jbc.M110.149880.

24. Richards S, Aziz N, Bale S, Bick D, Das S, Gastier-Foster J, et al. Standards and guidelines for the interpretation of sequence variants: a joint consensus recommendation of the American College of Medical Genetics and Genomics and the Association for Molecular Pathology. Genetics in Medicine. 2015;17(5):405–24. doi: 10.1038/gim.2015.30.

25. Whitfield TT, Sharpe CR, Wylie CC. Nonsense-Mediated mRNA Decay in Xenopus Oocytes and Embryos. Developmental Biology. 1994;165(2):731–4. doi: 10.1006/dbio.1994.1289.

26. Okiyoneda T, Harada K, Takeya M, Yamahira K, Wada I, Shuto T, et al. ΔF508 CFTR Pool in the Endoplasmic Reticulum Is Increased by Calnexin Overexpression. Molecular Biology of the Cell. 2004;15(2):563–74. doi: 10.1091/mbc.e03-06-0379.

27. Hibino H, Inanobe A, Furutani K, Murakami S, Findlay I, Kurachi Y. Inwardly Rectifying Potassium Channels: Their Structure, Function, and Physiological Roles. Physiological Reviews. 2010;90(1):291–366. doi: 10.1152/physrev.00021.2009.

28. Xu B, Levchenko V, Bohovyk R, Ahrari A, Geurts AM, Sency V, et al. Characterization of a novel variant in KCNJ16, encoding K(ir)5.1 channel. Physiol Rep. 2024;12(20):e70083. doi: 10.14814/phy2.70083.

29. Singhania P, Ponnusamy R, Dhar A, Agarwal A, Santra G. Distal Renal Tubular Acidosis: A Conundrum of Short Stature, Failure to Thrive, Rickets, and Nephrocalcinosis. Cureus. 2025. doi: 10.7759/cureus.90337.

30. Marcus DC, Wu T, Wangemann P, Kofuji P. KCNJ10 (Kir4.1) potassium channel knockout abolishes endocochlear potential. American Journal of Physiology-Cell Physiology. 2002;282(2):C403–C7. doi: 10.1152/ajpcell.00312.2001.

31. Burns JC, Kelly MC, Hoa M, Morell RJ, Kelley MW. Single-cell RNA-Seq resolves cellular complexity in sensory organs from the neonatal inner ear. Nature Communications. 2015;6(1). doi: 10.1038/ncomms9557.

32. Tanemoto M, Fujita A, Higashi K, Kurachi Y. PSD-95 Mediates Formation of a Functional Homomeric Kir5.1 Channel in the Brain. Neuron. 2002;34(3):387–97. doi: 10.1016/s0896-6273(02)00675-x.

33. Lv J, Fu X, Li Y, Hong G, Li P, Lin J, et al. Deletion of Kcnj16 in Mice Does Not Alter Auditory Function. Front Cell Dev Biol. 2021;9:630361. doi: 10.3389/fcell.2021.630361.

34. Konstas A-A, Korbmacher C, Tucker SJ. Identification of domains that control the heteromeric assembly of Kir5.1/Kir4.0 potassium channels. American Journal of Physiology-Cell Physiology. 2003;284(4):C910–C7. doi: 10.1152/ajpcell.00479.2002.

35. Blanchard A, Bockenhauer D, Bolignano D, Calò LA, Cosyns E, Devuyst O, et al. Gitelman syndrome: consensus and guidance from a Kidney Disease: Improving Global Outcomes (KDIGO) Controversies Conference. Kidney International. 2017;91(1):24–33. doi: 10.1016/j.kint.2016.09.046.

36. Seys E, Andrini O, Keck M, Mansour-Hendili L, Courand P-Y, Simian C, et al. Clinical and Genetic Spectrum of Bartter Syndrome Type 3. Journal of the American Society of Nephrology. 2017;28(8):2540–52. doi: 10.1681/asn.2016101057.

37. Juang J-MJ, Lu T-P, Lai L-C, Ho C-C, Liu Y-B, Tsai C-T, et al. Disease-Targeted Sequencing of Ion Channel Genes identifies de novo mutations in Patients with Non-Familial Brugada Syndrome. Scientific Reports. 2014;4(1). doi: 10.1038/srep06733.

