## Supplementary material for "Type, location and zygosity of *KCNJ16* mutations may determine the clinical severity of Hypokalemic Tubulopathy and Deafness (HkTD)": Supplemtal Figure 1

[1] Pediatric Nephrology Department, Cruces University Hospital, Barakaldo, Spain
[2] University of the Basque Country (EHU), Leioa, Bizkaia, Spain
[3] Biobizkaia Health Research Institute. Plaza de Cruces 12, 48903, Barakaldo, Bizkaia, Spain
[4] Department of Pediatric Nephrology, University Children's Hospital, Philipps University Marburg, Germany
[5] Biomedical Research Network Center for Diabetes and Associated Metabolic Diseases (Centro de Investigación Biomédica en Red de Diabetes y Enfermedades Metabólicas Asociadas, CIBERDEM), Biomedical Research Network Center for Rare Diseases (Centro de Investigación Biomédica en Red de Enfermedades Raras, CIBERER), EndoERN, Barakaldo, Bizkaia, Spain
[6] Pediatric Nephrology. Hospital Vall d'Hebron. University Autonomous Barcelona. Renal Physiopathology Group. Vall d`Hebron Research Institute. Ricords Network, Barcelona, Spain
[7] Pediatric Department. Hospitalario General de Fuerteventura. Puerto del Rosario, Las Palmas, Spain.
[8] Pediatric Nephrology Department. Complejo Hospitalario Universitario Insular-Materno Infantil. Las Palmas de Gran Canaria, Las Palmas, Spain.
[9] Nephrology Department. Hospital Universitario La Paz, Instituto de Investigación La Paz, IdiPAZ, Madrid, Spain
[10] Clinical Genetics Department. Complejo Hospitalario Universitario Insular-Materno Infantil. Las Palmas de Gran Canaria, Las Palmas, Spain
[11] Institute for physiology and pathophysiology, Philipps University Marburg, 35037, Marburg, Germany

* Shared Senior and Correspondence

**Correspondence:**

Dr. Aparna Renigunta
Department of Pediatric Nephrology,
University Children's Hospital, Philipps University Marburg,
35043, Marburg, Hessen, Germany
Ph: +49 64215862968


Dr. Leire Madariaga
Department of Pediatric Nephrology
Cruces University Hospital, University of the Basque Country, Biobizkaia Health Research Institute
Plaza de Cruces s/n
48903, Barakaldo, Bizkaia, Spain
Ph: +34946006329


### Supplementary Figure 1

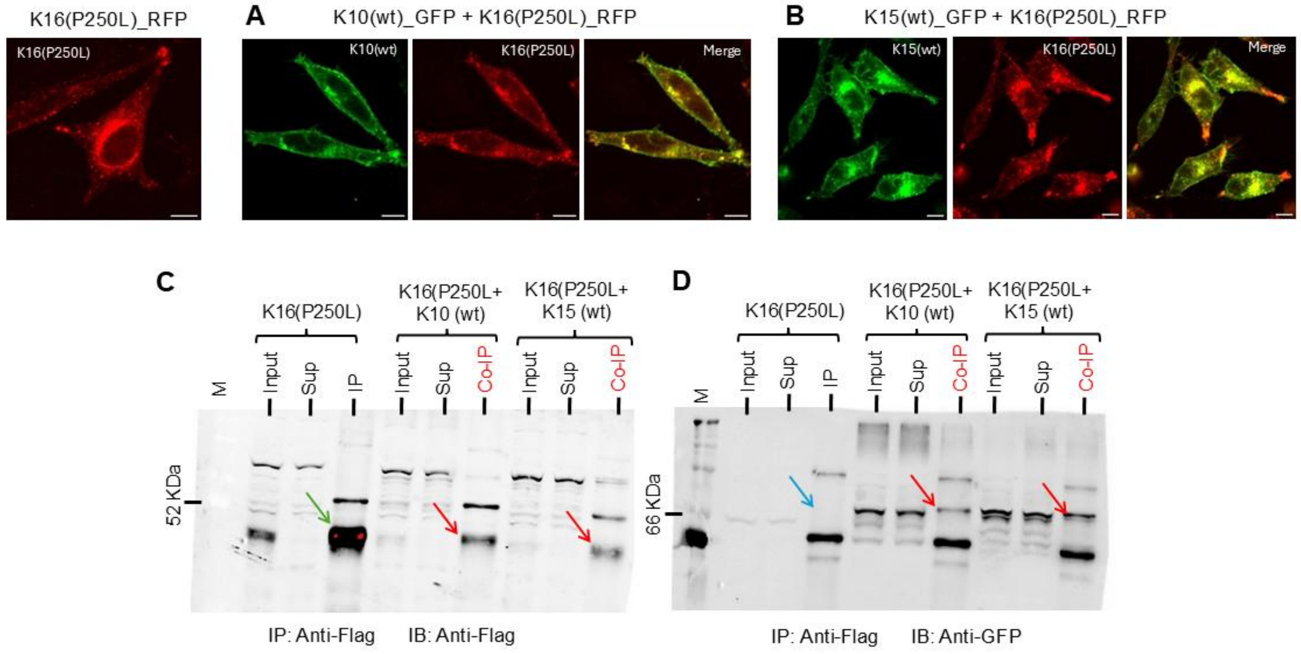


**SI: Fig.1** Membrane localization of K10 **(A)** and K15 **(B)** when co-expressen with K16(P250L). HeLa cells were co-transfected with GFP-tagged *KCNJ10* (green) or *KCNJ15* (green) together with RFP-tagged *KCNJ16*(P250L) (red). Overlaid images (Merge) highlight areas of co-localization in yellow. All images were acquired 48 hours post-transfection. Scale bar = 10 µm. **(C-D)** Effect of K16(P250L) on protein-protein interaction with K10 and K15. Co-immunoprecipitation experiments were performed in HEK293 cells to investigate the interaction between Flag-tagged K16(P250L) (K16-Flag, ~50 kDa) and either GFP-tagged wild-type K10 (K10-GFP, ~70 kDa) or K15 (K15-GFP, ~70 kDa). Cell lysates were prepared and subjected to immunoprecipitation using anti-Flag beads. Western blots were subsequently probed with anti-Flag antibody to detect K16(P250L)-Flag (green arrows in IP and red arrows in Co-IP fractions) and anti-GFP antibody to detect co-immunoprecipitated K10-GFP **(C)** or K15-GFP **(D)** (red arrows in Co-IP lanes). Control lanes (blue arrows) where the respective interacting partner (K10 or K15) was not co-expressed, were included to assess background binding. Lane designations across all panels include M (Marker), Input (total cell lysate), Sup (supernatant after IP), IP (immunoprecipitate), and Co-IP (co-immunoprecipitate).
